# Emergence of first strains of SARS-CoV-2 lineage B.1.1.7 in Romania

**DOI:** 10.1101/2021.01.29.21250643

**Authors:** Andrei Lobiuc, Mihai Dimian, Olga Sturdza, Roxana Filip, Mihai Covasa

## Abstract

United Kingdom reported the emergence of a new and highly transmissible SARS-CoV-2 variant B.1.1.7. that rapidly spread to other contries. The impact of this new mutation that occurs in the S protein, on infectivity, virulence and current vaccine effectiveness is still under evaluation. We have identified the first cases of the B.1.1.7 variant in samples collected from Romanian patients, of which one was traced to the UK region where the new variant was originally sequenced. Mutations in the Nsp3 protein, N844S and D455N and L15F in Orf3a were also detected, indicating common ancestry with UK strains as well as remote connections with strains from Nagasaki, Japan. These results indicate, for the first time, the presence and characteristics of the new variant B.1.1.7 in Romania and underscore the need for increased genomic sequencing in confirmed COVID-19 patients.

## Introduction

A new SARS-CoV-2 variant, with a N–Y substituion in the 501 position of the S (spike) protein, was detected in the UK in the fall of 2020. An initial version of the virus, termed 501 N, with fewer mutations, occurred in late September in Wales, followed by the current version (VUI-202012/01), giving rise to lineage B.1.1.7, which began to spread rapidly in the UK and then globally [1]. The new variant totals 18 particular mutations, of which several bare biological significance and are of epidemiological interest. Among most notable mutations is N501Y, within spike protein, which corresponds to the receptor bidning domain (RBD) of the virus, where attachment to host ACE2 enzyme takes place. Other important mutations are the deletion of two aminoacids, histidine and valine, at positions 69 and 70, and a substitution at position 681, within the same spike protein. Of great concern is the increased transmisibility and disease severity than older variants, raising questions on its potential avoidance of succesful nucleic acid amplification diagnostic or even vaccine effectiveness [2]. On January 8, 2021 Romania confirmed the first case of COVID 19 infection with the new strain, in a patient from Giurgiu, without a history of travel in the UK or contact with individuals from UK. On January 22, 2021, two additional individuals from Bucharest were identified with the new strain. They reported no travel history, good clinical condition, and were isolated at home under the supervision of family physician. A fourth case was reported in Suceava county, N-E of Romania, on January 25, 2021 in an indvidual who arrived from UK. A fifth reported case, at the time of writing this article, was confirmed on January 26, 2021 in a patient from Constanta, S-E of Romania, with no travel history or contact with individuals infected with the new strain. Considering the time of B.1.1.7 appearance in Europe, its fast spreading compared to earlier strains and the lack of genomic sequencing in Romania, there exists the possibility that the new variant is far more widespread in Romania. In this paper, we report the identification of the new B.1.1.7 SARS-CoV-2 variant in Romania and present its characteristics in sequenced samples with the aim of setting a further base for comparison of virus transmission.

## Materials and Methods

Twenty samples, collected from patients in the cities of Cluj, Craiova and Suceava counties from Romania were selected for analysis, including patients with possible contacts with UK infected individuals. Samples were quantified for viral titers and RNA amounts, through qPCR and Qubit methods, respectively. RNA extracts were reverse transcribed and libraries were prepared using Ampliseq SARS-CoV-2 primer panels and workflow. Automatic library templating was performed using Ion Chef equipment and sequencing was carried out on Ion GeneStudio S5 with Ion 540 chips. Sequencing reads and assemblies were checked for quality using Ion Torrent Suite software plugins. Aminoacid substitution analysis was performed using CoV-Glue platform (http://cov-glue.cvr.gla.ac.uk/#/replacement). The B.1.1.7 SARS-CoV-2 sequence was uploaded into GISAID, under ID EPI_ISL_869241. Consensus sequence and available Romanian sequences, from different laboratories, belonging to clade B.1.1.7 in Romania, in GISAID were aligned to reference strain using MAFFT algorithm and maximum likelihood trees were obtained with MegaX software.

## Results and Discussions

Among the 20 samples sequenced, one presented characteristic mutations of the B.1.1.7. SARS-CoV-2 variant. Phylogenetic placement of this sample and the other from Romania within the same lineage, included in GISAID, shows the clear distinction of this lineage from the early, 2020 strains, including the ones from England and Wales (Fig. 1).

**Figure 1.**
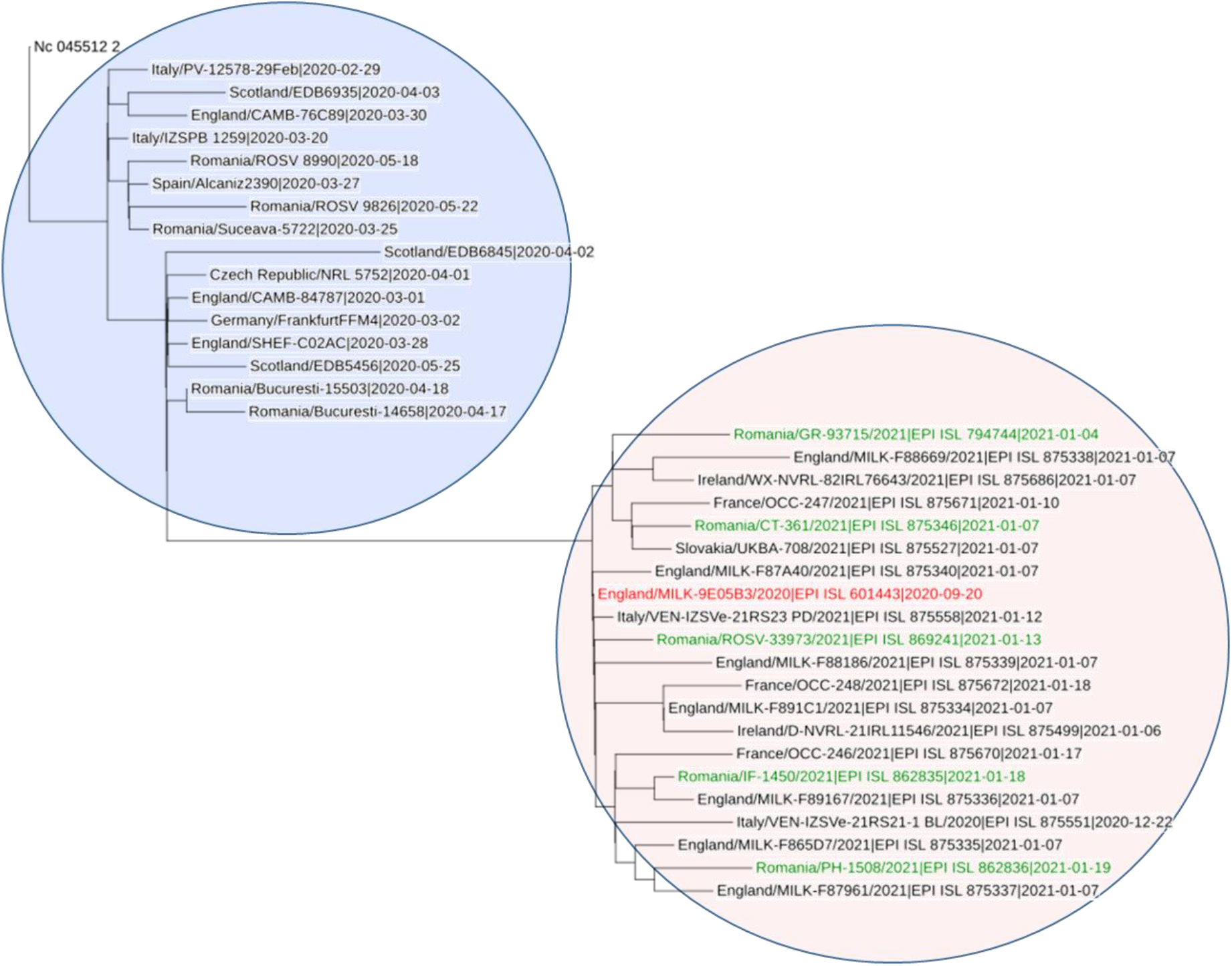
Phylogenetic placement of pre-B.1.1.7 samples (blue area) and B.1.1.7 samples (red area), from different European countries, including Romanian strains (green text).

Considering all Romanian GISAID accessions belonging to this clade, a synopsis of all mutations was constructed (Table 1). All Romanian samples share all 18 mutations, characteristic of B.1.1.7 strain, however some of them bare additional ones.

**Table 1.** List of mutations encountered in Romanian B.1.1.7 samples (green entries indicate mutations characteristic to B.1.1.7, bold entries indicate singular, non-B.1.1.7, mutations among Romanian samples)

| Genome region | Orig . AA | Pos. | Subs . AA | EPI_ISL_8 69241 (SV) | EPI_ISL_7 94744 (GR) | EPI_ISL_8 62835 (IF) | EPI_ISL_8 62836 (PH) | EPI_ISL_8 75346 (CT) |
| --- | --- | --- | --- | --- | --- | --- | --- | --- |
| S | N | 501 | Y | yes | yes | yes | yes | yes |
|  | A | 570 | D | yes | yes | yes | yes | yes |
|  | D | 614 | G | yes | yes | yes | yes | yes |
|  | P | 681 | H | yes | yes | yes | yes | yes |
|  | T | 716 | I | yes | yes | yes | yes | yes |
|  | S | 982 | A | yes | yes | yes | yes | yes |
|  | D | 1118 | H | yes | yes | yes | yes | yes |
|  | HV | 69-70 | del | yes | yes | yes | yes | yes |
|  | Y | 144 | del | Yes | yes | yes | Yes | Yes |
| N | D | 3 | L | Yes | yes | yes | Yes | yes |
|  | R | 203 | K | Yes | yes | yes | Yes | yes |
|  | G | 204 | R | Yes | yes | yes | Yes | yes |
|  | S | 235 | F | Yes | yes | yes | Yes | yes |
| nsp 2<br>(ORF1ab) | A | 306<br>(486) | V |  |  |  |  | Yes |
| nsp3<br>(ORF1ab) | T | 183<br>(1001) | I | Yes | yes | yes | Yes | yes |
|  | N | 844<br>(1662) | S | Yes |  |  |  |  |
|  | A | 890<br>(1708) | D | Yes | yes | yes | Yes | yes |
|  | I | 1412<br>(2230) | T | Yes | yes | yes | Yes | yes |
|  | D | 455 | N |  |  |  | Yes |  |
| nsp4<br>(ORF1ab) | F | 17<br>(2780) | L |  |  |  | Yes |  |
| nsp6<br>(ORF1ab) | SF | 106-108<br>(3675-3677) | del | Yes | yes | yes | Yes | yes |
| nsp12<br>(ORF1ab) | K | 160<br>(4552) | N |  |  |  |  | yes |
|  | P | 323<br>(4715) | L | Yes | yes | yes | Yes | yes |
| nsp13<br>(ORF1ab) | K | 460<br>(5748) | R |  |  | yes | Yes |  |
| nsp14<br>(ORF1ab) | A | 119<br>(6044) | V |  | yes |  |  |  |
|  | E | 347<br>(6272) | G |  |  | yes |  |  |
| nsp15<br>(ORF1ab) | P | 111<br>(6563) | T |  | yes |  |  |  |
| ORF 3a | L | 15 | F |  |  |  | Yes |  |
| ORF 7a | Y | 20 | N |  |  |  | Yes |  |
| ORF 8 | Q | 18 | stop | Yes |  |  |  |  |
|  | Q | 27 | stop | Yes | yes | yes | Yes | yes |
|  | R | 52 | I | Yes | yes | yes | Yes | yes |
|  | Y | 73 | C | Yes | yes | yes | Yes | yes |
|  | K | 68 | stop |  | yes |  |  | yes |

*Abbreviations:* SV, Suceava; GR, Giurgiu; IF, Ilfov; PH, Prahova; CT, Constanta

One such mutation is present only in the sample originating from Suceava (SV), affecting ORF8 protein, where a stop codon is gained, by changing C to T nucleotides in position 27945 in the genome. This mutation has been already encountered, according to GISAID, in over 500 samples from April to September 2020. Of these, 73% belong to UK collected specimens [3]. A second ORF8 truncation, not currently described for B.1.1.7 strains, appears in the samples from Giurgiu (GR) and Constanta (CT), in position 68, also gaining a stop codon. Previous occurences of this mutation are seen in 120 samples from GISAID, of which 90% are from UK and 36% originate from Milton Keynes laboratories, where the original B.1.1.7 strain was sequenced [4]. Such mutations indicate that, although B.1.1.7 originates in UK, the set of characteristic viral alterations appeared much earlier and were grafted onto several already circulating different strains in the region. This idea is supported by the fact that, although the first sequenced samples carrying the new strain originated in Kent and Greater London, on 20^th^ and 21^st^ of September 2020, respectively, [5], the hallmark N501Y mutation first appeared in Italy, in August 2020 [6]. However, at this point, the Romanian strains bearing the particular ORF8 mutations described above clearly originated in the UK, which is also supported by the fact that the patient from Suceava county arrived in Romania from the UK.

Strains without a functional ORF8 protein are considered to present epitope loss, which may decrease the accuracy of serological testing, whereas ORF8 antibodies could offer information on both acute and convalescent antibody response. Furthermore, ORF8 truncated proteins decrease disease severity and asymptomatic or mildly diseased carriers might not be detected [7]. As such, the significance of ORF8 truncations in the context of B.1.1.7 strains should be promptly investigated, considering that mutations in S gene characteristic to this lineage, particularly the deletion at positions 69-70, may elude PCR diagnostic with certain kits, which have been used in the UK for a while [8]. This type of behaviour could be indirectly, but significantly, linked to increased transmissibility of the virus, as potentially infected population carrying this strain might have not been accurately identified as such.

Another noteworthy mutation is N844S within Nsp3 protein present in SV sample, which is recorded only in other 8 samples sequenced so far, most of them also from England [9]. The sample from Prahova (PH) presents also a mutation in Nsp3, D455N, which is recorded in only 1 other sample from Japan [10], from April 2020, belonging to clade B1.1. The PH sample is again distinct from others in Romania through the appearance of L15F in Orf3a, a mutation recorded in 5 samples from Nagasaki, Japan, sampled in April 2020, among 243 samples worldwide, mostly from UK [11]. Although Japan samples do not belong to B.1.1.7 lineage, the coincidental presence of these mutations might indicate ancestry in PH sample. Other individual mutations in GR and IF samples are common with UK sampled strains. The CT sample displays two additional mutations, not encountered in other Romanian samples. The first, in the non-structural protein 2 (nsp2), changing A to V in position 306, mutation seen in other 209 GISAID samples. Out of these, besides UK, certain percentages belong to Norway, Denmark, USA and Belgium [12] (CoV-Glue, 2021f). A second particular mutation is in nsp12, changing K to N in position 160, encountered in other 27 samples, including the ones from USA, Italy and Scotland [13].

At the moment, there are over 32,500 B.1.1.7 accessions deposited in GISAID, out of which aproximately 30,000 are from UK and 5 are from Romania. This lineage is of major interest, due to the fact that three of its mutations might contribute to higher infectivity and transmisibility. Namely, the N50Y mutation of S gene significantly increases the force and number of interactions with the human receptor ACE2 [14,15]. The two aminoacid deletion at positions 69-70 in the same S gene leads to systematically biased diagnostic tests and doubles the reproductive advantage and numbers of the virus [16]. Furthermore, the P681H mutation of S protein might influence the cleavage of S protein through effest on S1/S2 subunits furin cleavage site [17]. Identification of new mutations is crucial in designing correct diagnostic reagents [18], slowing transmission and vaccine reconfiguration against new variants. Also, particular mutations, besides those specific to B.1.1.7, may, in the future, aid in tracing virus movements across Romania and worldwide. Many European countries, including Romania, lag in genomic sequencing and EU recommends increased focused sequencing based on epidemiological data, transmission rates, infectivity, treatment failure and S-gene “drop-out” in PCR testing. Therefore, thorough characterisation of strains circulating in Romania also contributes to developing useable diagnostic tests and vaccines, especially in the light of notable differences between strains belonging to the same clade and the evolutionary capacity of SARS-CoV-2.

## Data Availability

All data are included in the paper. GISAID accession ID EPI_ISL_869241

## Authors’ contributions

AL, MD and MC contributed to the conception and design of the study. AL, OS, RF contributed to acquisition of data. AL and MC drafted the article. All authors revised the article for important intelectual content and aproved the final version of the manuscript.

## Ethics approval and consent to participate

The study was aproved by the ethics committee of University Stefan cel Mare of Suceava, Romania (no. 11733/14.07.2020) and of Suceava County Emergency Hospital (no.

17669/13.07.2020). All participants signed individual informed consent.

